# Incidence and Risk Factors of Mortality in Adults with Congenital Heart Disease: Results from the Mayo Adult Congenital Heart Disease Registry

**DOI:** 10.64898/2026.08.07.26359991

**Authors:** Naveenkumar Nallathambi, Ishika Gupta, Keerthika Vijayakumar, William R Miranda, Alexander Egbe, Luke J. Burchill, Brian D. Lahr, Alexander T. Lee, Abhishek Deshmukh, Samuel J. Asirvatham, Malini Madhavan

## Abstract

**Background:** Adults with congenital heart disease (ACHD) represent a rapidly expanding population with evolving mortality patterns. Despite improved survival, excess mortality persists.

**Objective:** To evaluate the incidence, causes, and predictors of mortality in a contemporary ACHD cohort.

**Methods:** We performed a retrospective cohort study of adults (≥18 years) first evaluated at Mayo Clinic from 2002–2023. Baseline clinical, imaging, and electrocardiographic data were analyzed. Vital status was determined using institutional records and the Accurint national mortality database. Kaplan-Meier analysis and Cox proportional hazard models were used to evaluate mortality and identify independent predictors of mortality

**Results:** A total of 7,678 ACHD patients were included, with median age of 36.8 years and median follow-up of 11.4 years. During 78,768 patient-years of follow-up, 1,116 patients died (median age at death 57.2 years), corresponding to an annual mortality rate of 1.4%. The cumulative rate of all-cause mortality at 5, 10, 15, and 20 years was 6.9%, 12.0%, 19.0%, and 26.2%, respectively. When stratified by CHD complexity, the annual death rate in patients with severe CHD (2.4%/year) was twice that of patients with moderate or mild CHD (both 1.2%/year). Older age and ACHD subtypes, specifically, cyanotic heart disease (HR 3.9, 95% CI 2.9–5.3) and Fontan physiology (HR 3.2, 95% CI 2.3–4.4), were strongly associated with increased mortality. Additional independent predictors included male sex, ventricular dysfunction, advanced NYHA class, prior heart failure hospitalization, hypertension, smoking, coronary artery disease, renal dysfunction, and abnormal hemoglobin levels. Cardiovascular causes accounted for 57.7% of deaths with known etiology, predominantly heart failure (48.9%) and sudden cardiac death (21.9%), while non-cardiovascular causes were driven mainly by infection and malignancy.

**Conclusions:** In this large contemporary ACHD cohort, mortality was driven by ventricular dysfunction, heart failure, and systemic end-organ involvement in addition to the underlying congenital anatomy. Both cardiovascular and non-cardiovascular causes contributed significantly to mortality. These findings underscore the need for comprehensive multidisciplinary ACHD care focused on early recognition of cardiac functional decline, management of acquired comorbidities, and end-organ dysfunction.

## Introduction

Advances in pediatric cardiology, cardiac surgery, and critical care have transformed congenital heart disease (CHD) from a predominantly fatal condition of childhood into a chronic condition of adulthood (1). While survival has improved dramatically, longevity has not translated into normalcy — ACHD patients face a lifetime of residual hemodynamic burden, recurrent interventions, and progressive multiorgan dysfunction that collectively diminish long-term prognosis. Heart failure and arrhythmia remain the most recognized drivers of morbidity and mortality in this population (2). However, as the ACHD population ages, acquired cardiovascular comorbidities — coronary artery disease (CAD), hypertension, obesity — and non-cardiovascular conditions such as malignancy are emerging as increasingly important contributors to premature death.

Understanding the full spectrum of mortality in contemporary ACHD patients is therefore essential not only to guide individualized risk stratification and surveillance, but to inform the design of ACHD care models that extend beyond traditional cardiac-focused follow-up. Most existing mortality data derive from earlier surgical eras, younger cohorts, or population-based registries that may underrepresent the high-risk, complex end of the ACHD spectrum seen at tertiary referral centers. Contemporary, granular data on the incidence, causes, and predictors of death — including the growing contribution of non-cardiovascular mortality — remain limited. This study addresses that gap by evaluating mortality in a large, contemporary cohort of ACHD patients followed at a major referral center over two decades.

## Methods

### Study design and population

This is a retrospective cohort study of adult patients aged ≥18 years with CHD first evaluated at Mayo Clinic between 2002 and 2023 in the Mayo adult congenital heart disease registry. The study was approved by the Mayo Clinic Institutional Review Board and informed consent was waived due to minimal risk. Baseline demographic, clinical, laboratory, electrocardiographic, echocardiographic and other disease-specific variables were extracted from electronic medical records at the first clinical visit as adults (≥ 18 years of age). Patients were classified according to anatomic lesion severity into mild, moderate and severe ACHD using the Bethesda classification (3). The cohort was also classified into 7 broad ACHD subgroups: (1) right heart/conotruncal lesions (2) left heart disease (3) shunts (4) unrepaired cyanotic heart disease (5) other CHD with biventricular repair and systemic LV (6) systemic right ventricle and (7) Fontan palliation. The disease states included in each ACHD subgroup is presented in **Supplementary Table 1**.

Vital status was determined through linkage with the Accruint national mortality database, supplemented by institutional records. The primary cause of death was defined as the underlying disease or condition that directly led to the death as determined from the electronic health record. We classified deaths as cardiovascular (CV) and non-cardiovascular (non-CV). CV death was defined as death resulting from cardiac or vascular conditions, including heart failure, sudden cardiac death, post–cardiac surgical or procedural complications, myocardial infarction, stroke of cardiovascular origin, pulmonary embolism, vascular rupture, and infective endocarditis. Non-CV death was defined as death attributable to non-cardiac conditions such as infection or sepsis, malignancy, respiratory failure, renal or hepatic failure, gastrointestinal or other bleeding, trauma, neurologic disease, multi-organ failure, or complications of non-cardiac surgery. Sudden cardiac death was defined as an unexpected death due to presumed cardiac causes occurring within one hour of symptom onset, or an unwitnessed death in a previously stable individual without an identifiable non-cardiac cause.

### Statistical analysis

Survival time was measured from the date of first cardiovascular visit, with separate endpoints defined for death (from all causes), cardiovascular death, and sudden cardiac death. In the primary endpoint analysis, time until death was analyzed using the Kaplan-Meier and Cox proportional hazards (PH) regression methods, censoring the survival time for those patients alive at last follow-up. To conduct a detailed multivariable examination of baseline risk factors, an exhaustive list of candidate predictors was chosen a priori based on clinical relevance. After removing some variables from consideration due to extensive missing data or extremely low prevalence, a total of 34 predictor variables were entered into the Cox regression model. Multiple imputation (repeated 5 times) was used to fill in missing predictor data to allow their inclusion in the final analysis. Continuous predictors were modeled without assuming linearity using 4-knot restrictive cubic splines. Model results are presented as adjusted hazard ratios (HRs) with 95% confidence intervals (CIs), with the HRs for continuous variables computed at different predictor settings by choosing multiple values for comparison against a reference value. For example, we examined the relationship between body mass index (BMI) and survival time by estimating HRs from the model when BMI equals 20, 30 and 40 kg/m^2^ as compared to the reference value of 25 kg/m^2^.

For the secondary endpoint, time until cardiovascular death, cumulative mortality rates were estimated using a nonparametric cumulative incidence function estimator that accounted for the competing risk of death from non-cardiovascular or unknown causes. To elucidate risk factors of cardiovascular mortality, the multivariable regression analysis was repeated using the cause-specific hazard modeling approach. This technique fits a Cox PH regression model to the cardiovascular death endpoint with non-cardiovascular death considered a censoring event. Because cause of death information was frequently not available, we handled the deaths from unknown causes in this modeling using two strategies: one that coded these deaths as an additional censoring event (primary analysis) and one that coded the endpoint as missing thereby excluding these patients (sensitivity analysis). To reduce the candidate predictor variables from the prior multivariable model to a number that a model for cardiovascular mortality could reasonably support, we assumed a simpler functional form for each of the continuous variables by modeling them with one less knot in the spline function. An exception was ejection fraction, which was modeled assuming linearity in its effect after truncating high values at 60 (i.e., assumes a value above 60 has no incremental benefit). We also modeled NYHA as a continuous predictor assuming linearity rather than as a categorical variable. Additionally, we used a composite variable for any prior valve replacement rather than separate left-sided and right-sided valve replacement variables, and we combined the moderate/severe regurgitation variables into one variable, regardless of the valve (aortic, mitral, or tricuspid) affected. Finally, we dropped OSA, anti-pulmonary hypertensives, and MRA from the model due to the lower prevalence and priority of these variables. These data reduction actions resulted in a savings of 14 degrees of freedom. Results from this multivariable model are reported as adjusted cause-specific hazard ratios (cHRs) with 95% CIs.

As an exploratory analysis, the endpoint of sudden cardiac death was analyzed in similar fashion as cardiovascular death, except the risk factor analysis was limited to univariable regression analyses on a short pre-specified list of candidate risk factors given the relatively low number of sudden cardiac deaths. All analyses were done using R software version 4.4.1 (www.R-project.org). Statistical significance was defined by a p-value <0.05.

## Results

A total of 7,678 ACHD patients were included in the analysis, with median age at first evaluation of 36.8 (IQR 25.5 - 50.1) years and 48.6% were male. The baseline characteristics of the cohort are presented in **Table 1**. The distribution of ACHD subgroups included right-sided/conotruncal lesions as the largest subgroup (32.6%), followed by shunt lesions (29.6%), left-sided lesions (21.8%), systemic RV (6.1%), Fontan palliation (5.5%), cyanotic heart disease (2.4%), and other CHD with biventricular repair and systemic LV (2.0%). Overall, 24.5% of patients had mild (simple) CHD, 58.1% had moderate CHD, and 17.4% had severe (complex) CHD, as seen in **Supplementary Table 1**.

**Table 1.**
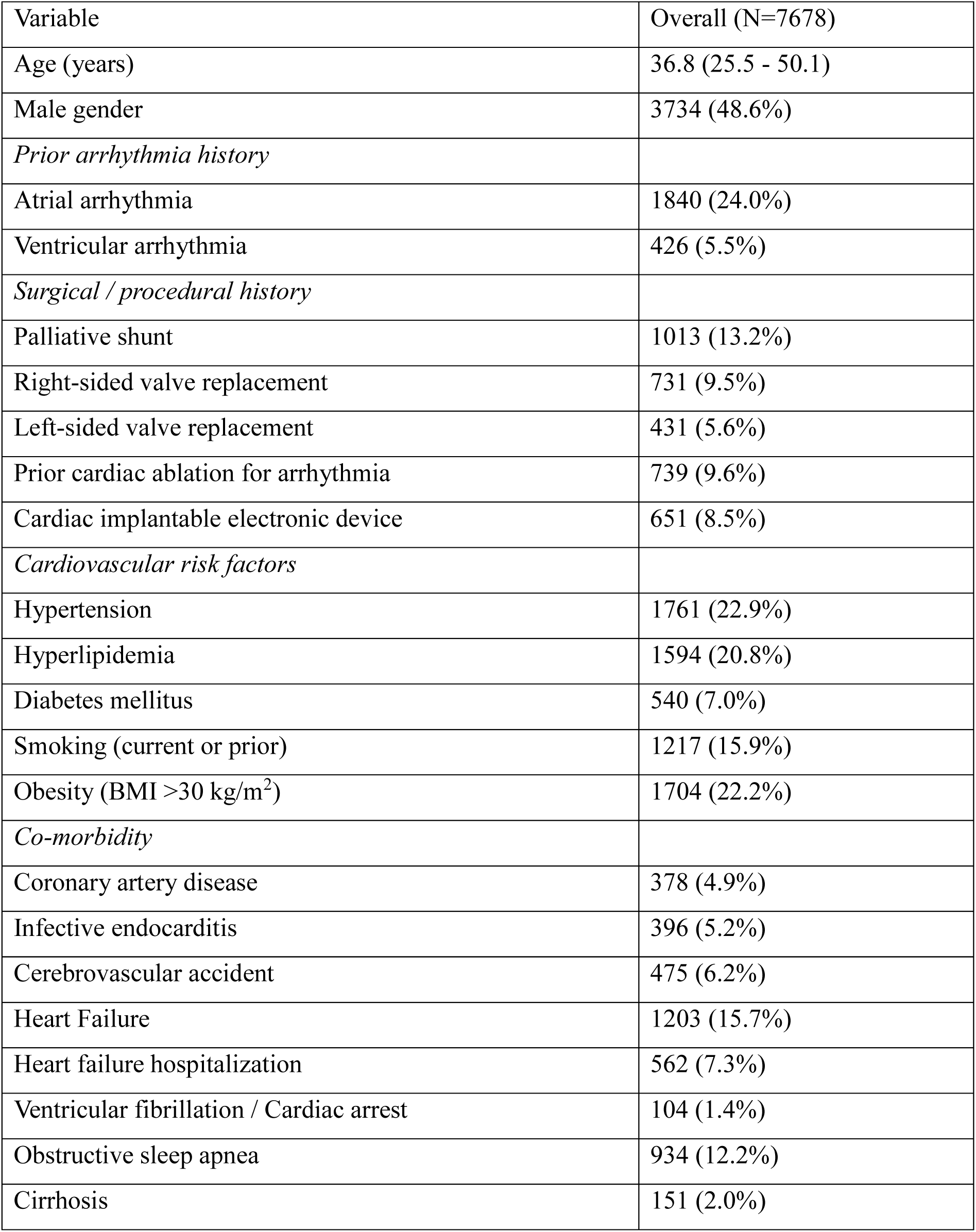
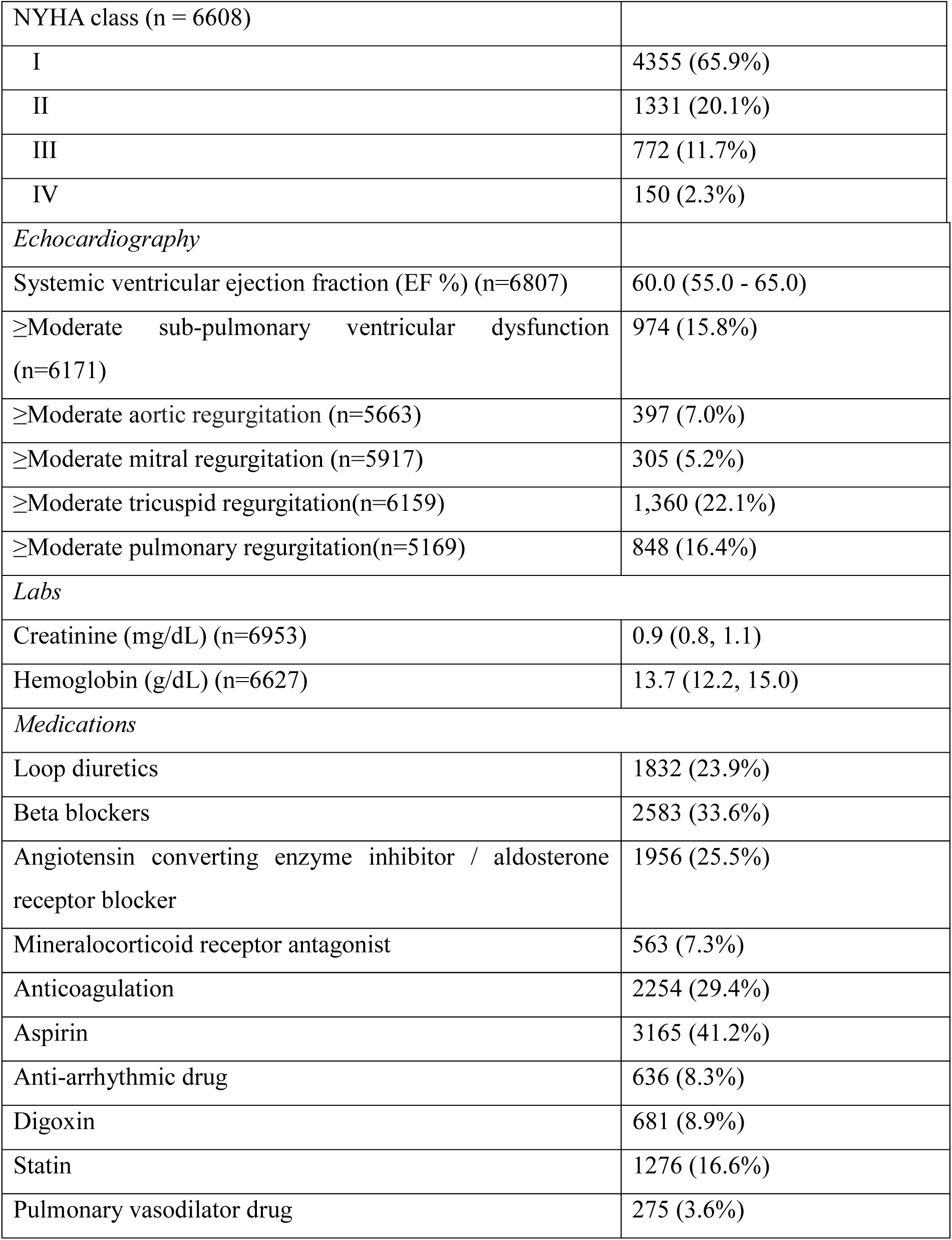
Baseline characteristics of the cohort of adults with congenital heart disease.

### All-cause mortality

Survival data were available on 78,768 cumulative patient-years collected during a median follow-up of 11.4 years (IQR 6.1-16.6). A total of 1,116 patients died (median age at death 57.2 years [IQR 41.1-70.7]), corresponding to an annual mortality rate of 1.4%. The cumulative rate of all-cause mortality at 5, 10, 15, and 20 years of follow-up was 6.9%, 12.0%, 19.0%, and 26.2%, respectively, according to the Kaplan Meier curve in **Figure 1**. When stratified by ACHD severity group, the annual death rate in patients with severe CHD was 2.4%/year, which is two-fold higher than the death rates in patients with moderate or mild CHD (both 1.2%/year). Median age at death was 70.6 years (IQR 57.1–80.7) for mild CHD, 60.2 years (IQR 45.2–70.8) for moderate CHD, and 41.7 years (IQR 34.3–55.2) for severe CHD.

**Figure 1.**
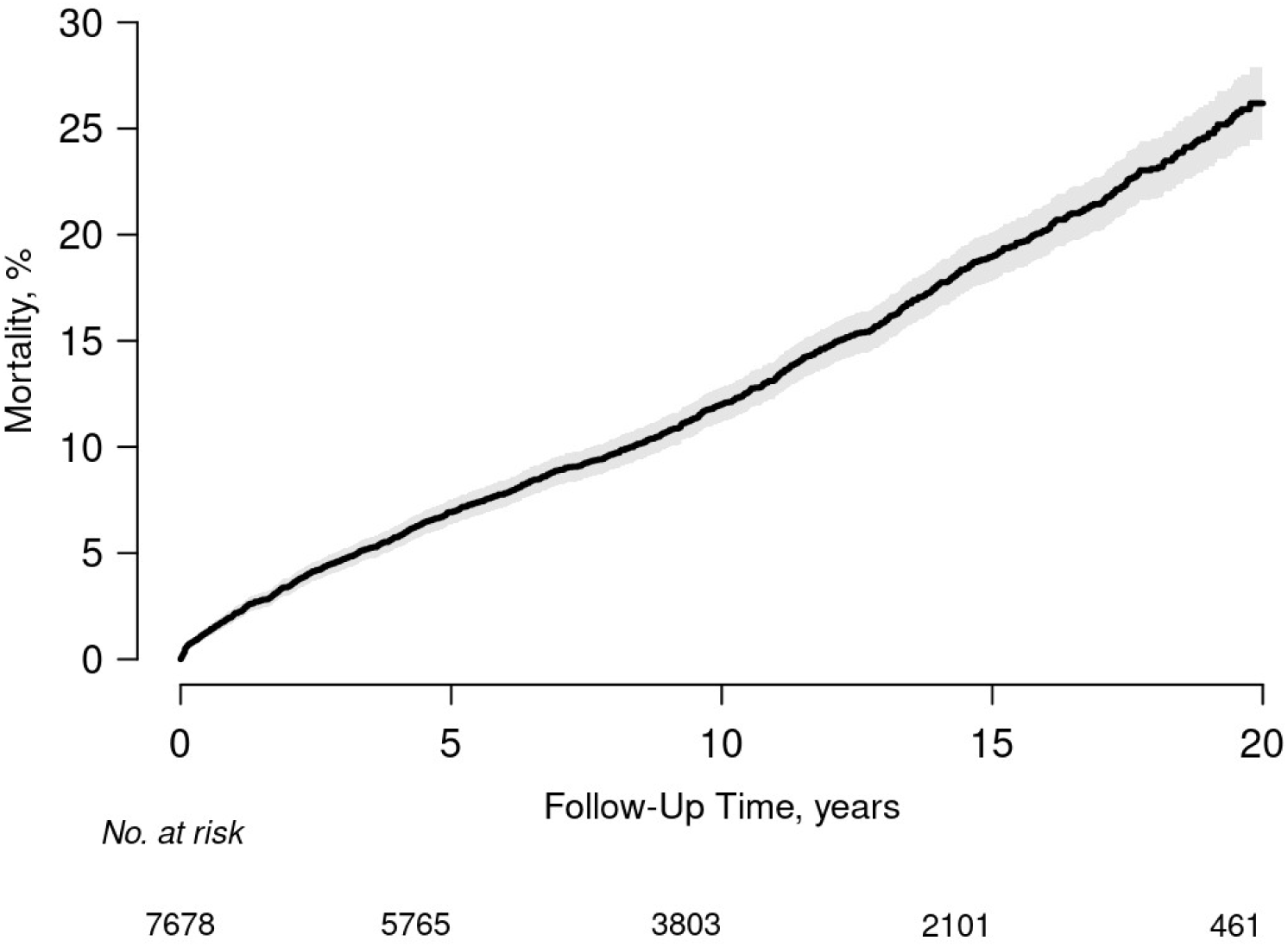
Kaplan–Meier analysis of time to all-cause mortality in the overall ACHD cohort.

The associations of potential predictors of all-cause mortality in the multivariable Cox regression analysis are presented in **Table 2**. The importance of each variable from the multivariable model is displayed using Wald χ² statistics adjusted for degrees of freedom (χ² − df), with larger values indicating greater independent contributions to the model as shown in **Figure 2**. Age and ACHD subgroup were the two strongest predictors of mortality. Compared with right-sided or conotruncal lesions, cyanotic heart disease and Fontan physiology were associated with higher mortality, whereas the risk from left-sided lesions, shunt lesions, systemic right ventricle, and other CHD subtypes was not significantly different. Several markers of ventricular dusfunction were independently associated with mortality, including systemic and sub-pulmonary ventricular dysfunction, advanced New York Heart Association (NYHA) functional class and prior heart failure hospitalization. Atrial and ventricular arrhythmias (composite of sustained and nonsustained ventricular tachycardia) were not associated with increased risk of mortality, nor was the severity of atrioventricular or semilunar valve regurgitation.

**Figure 2.**
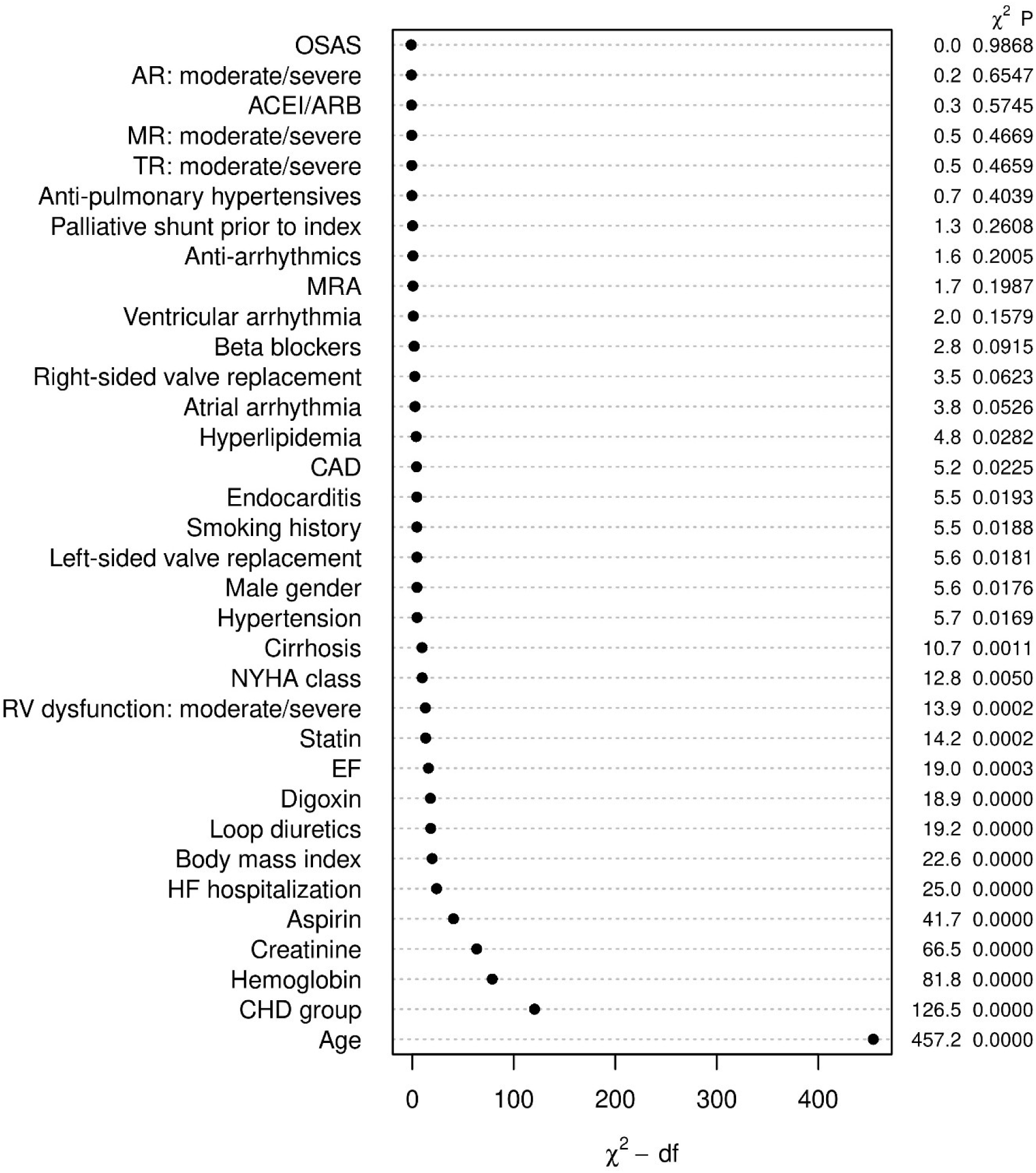
Relative importance of variables in the multivariable model, presented as adjusted Wald χ² statistics and corresponding *P* values.

**Table 2.**
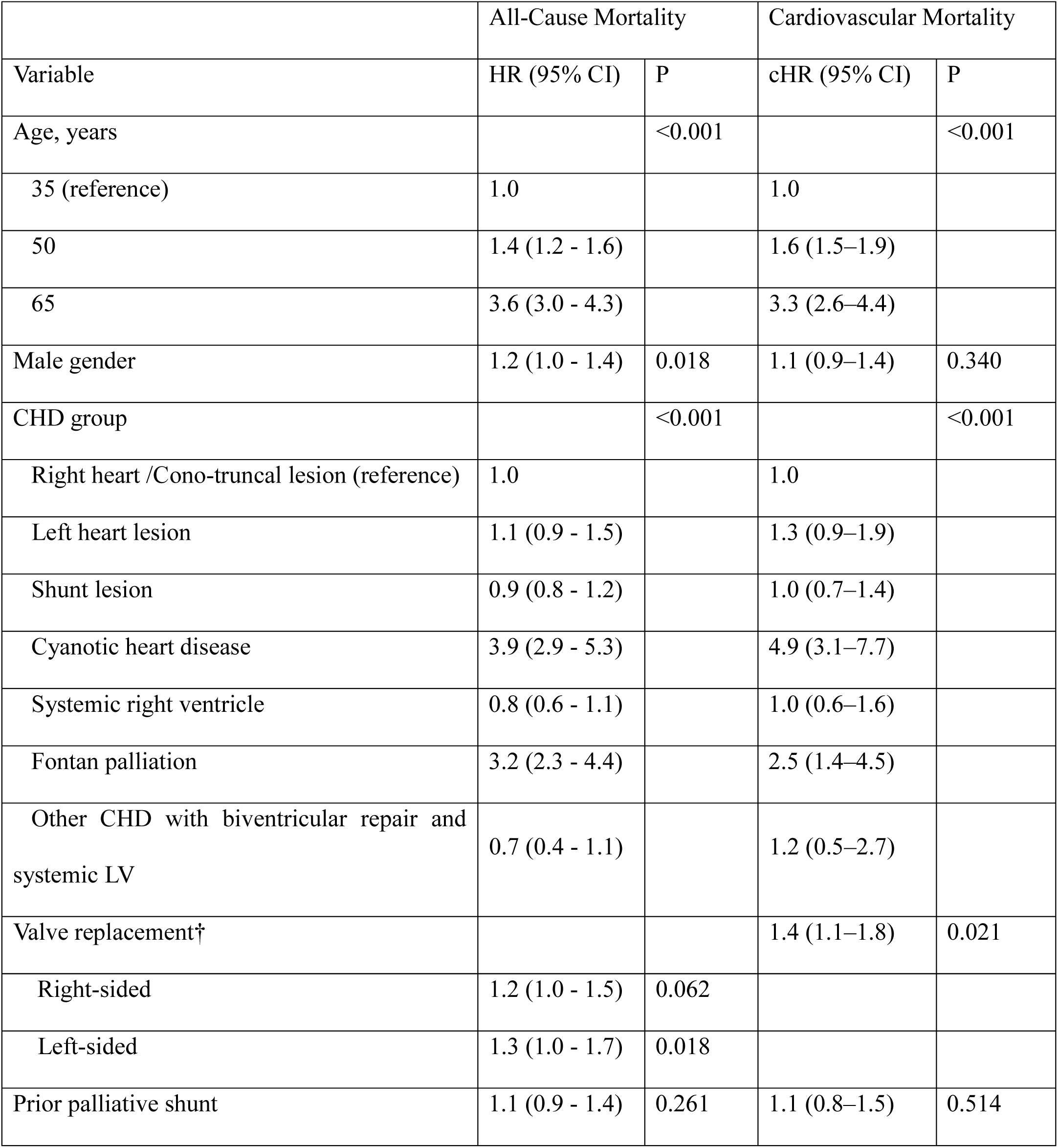
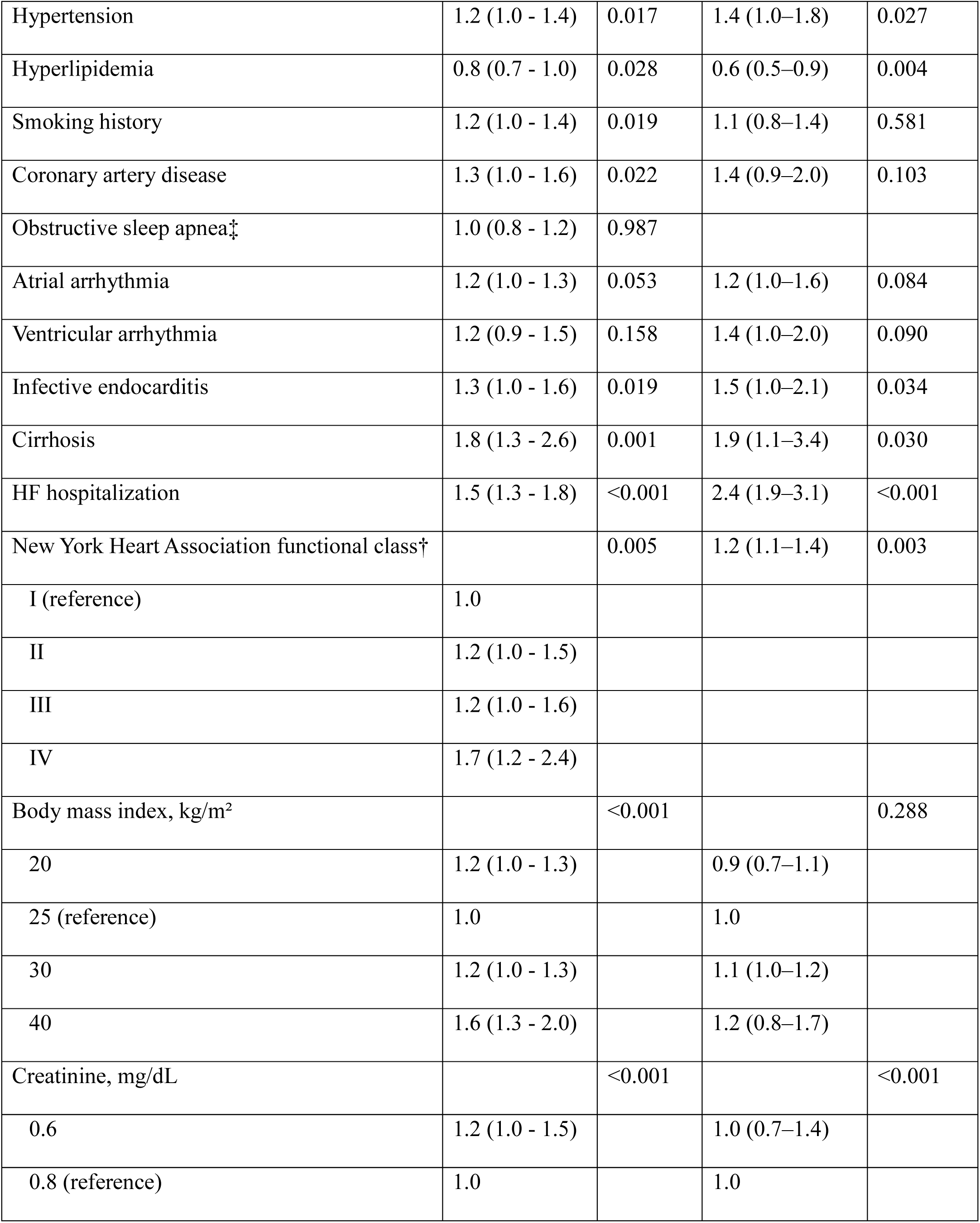
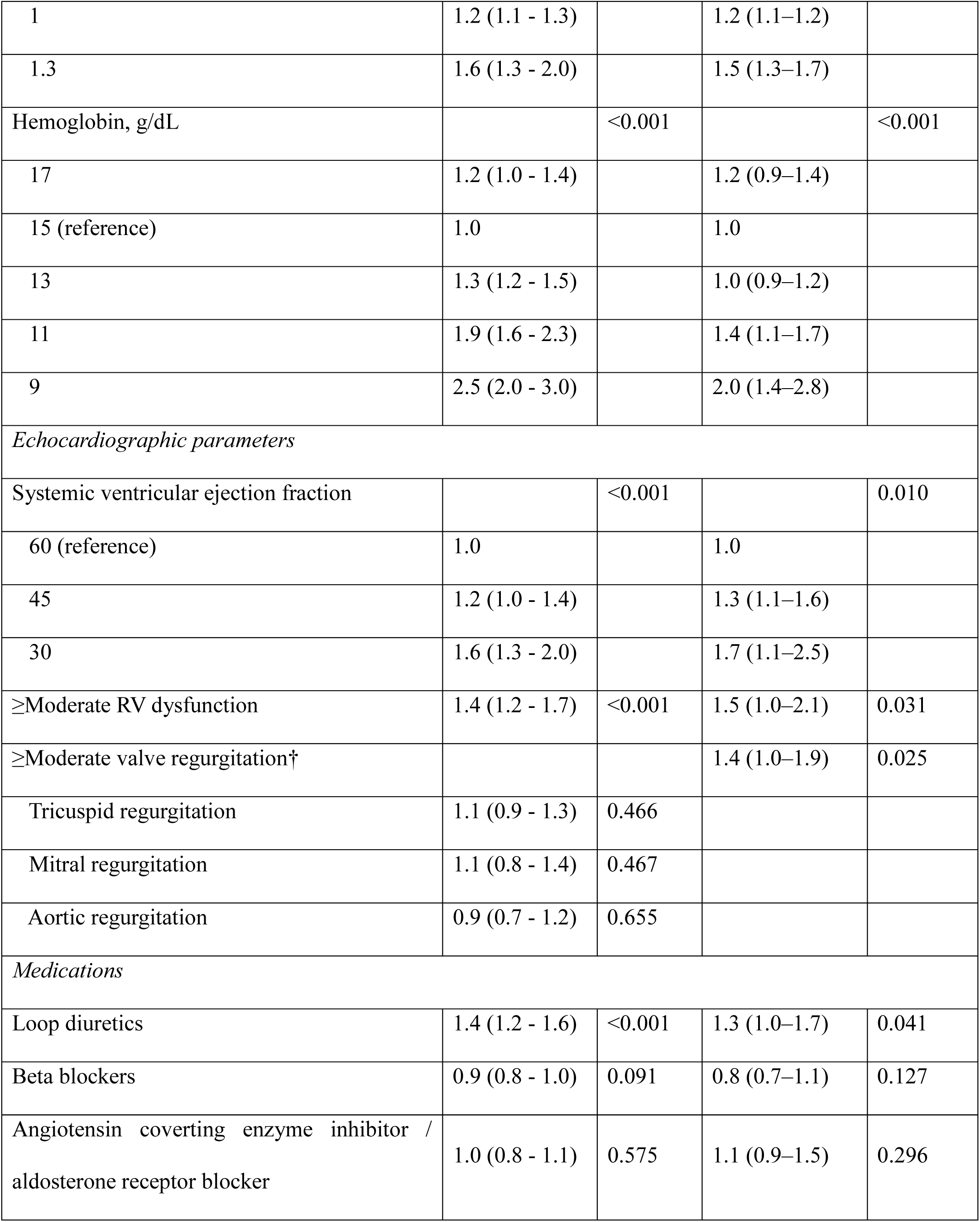
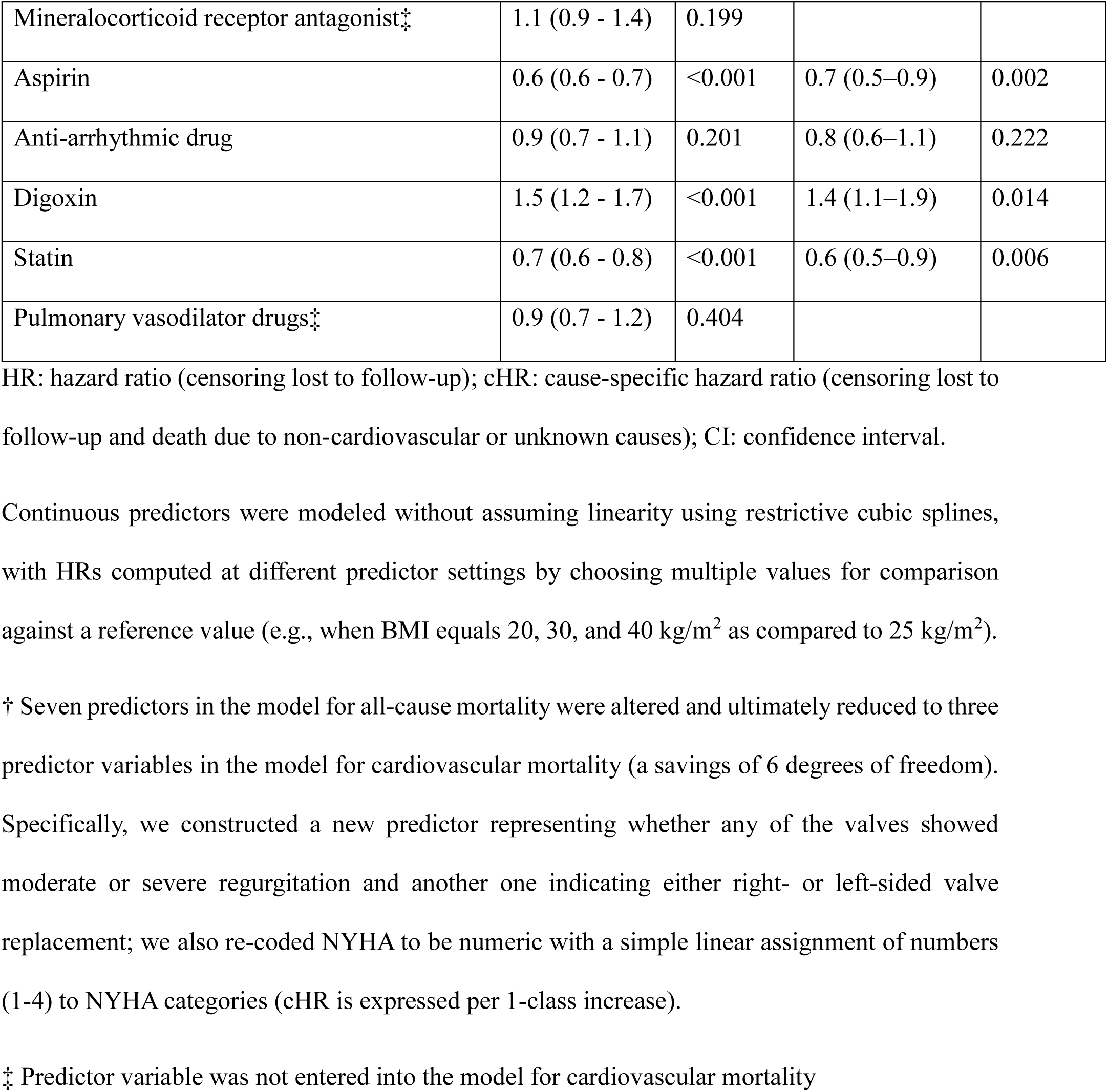
Multivariable Cox proportional hazards regression models for all-cause and cardiovascular-specific mortality.

Traditional cardiovascular risk factors such as hypertension and smoking as well as CAD were associated with increased mortality. Use of loop diuretics and digoxin were also associated with increased mortality, while aspirin and statin therapy were associated with lower mortality risk.

Continuous predictors were modeled using restricted cubic splines and the partial effects for each variable are plotted against the estimated log relative hazard of mortality in **Figure 3**. Age demonstrated a strong, monotonic association with increasing risk across the observed range. Hemoglobin and BMI exhibited a nonlinear, U-shaped relationship, with higher hazard at both extremes of values. Serum creatinine also had a non-monotonic relationship with mortality, with higher risk at very low values as well as at modestly elevated risk at higher values (i.e., > 1.0 mg/dL). Left ventricular ejection fraction was inversely and approximately linearly associated with the outcome.

**Figure 3.**
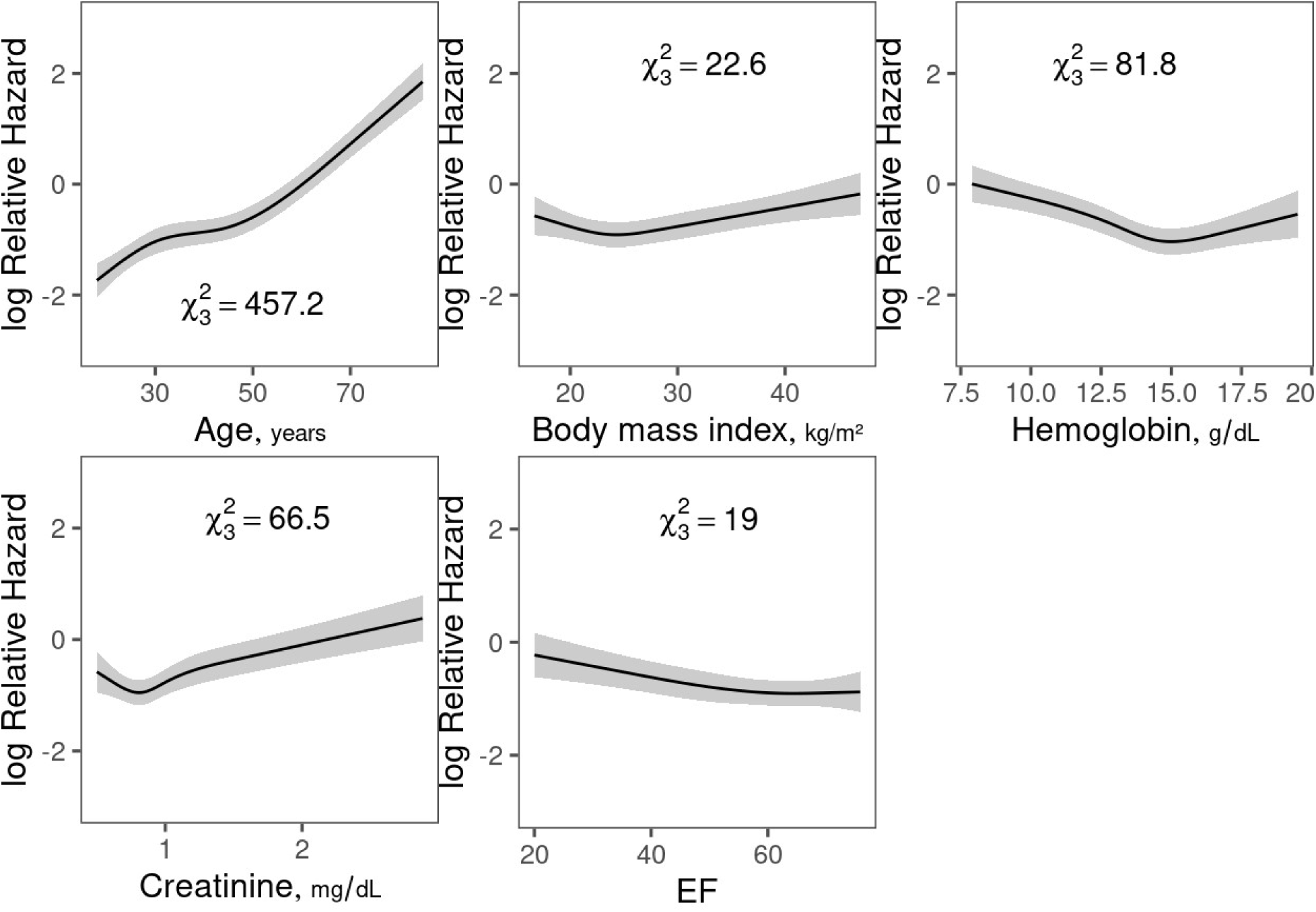
Relative hazard of all-cause mortality across the spectrum of age, body mass index, hemoglobin, creatinine, and systemic ventricular ejection fraction (EF).

### Cause of death

Of the 1,116 patients who died, deaths were cardiovascular-related in 370 (33.2%), not cardiovascular-related in 271 (24.3%), and of unknown cause in 475 (42.6%) patients. After taking the competing risk of earlier mortality into account, the overall estimates for cumulative CV death at 5, 10, 15, and 20 years of follow-up were 2.8%, 4.2%, 5.9%, and 7.6%, respectively. The corresponding rates of non-CV mortality were 1.8%, 3.1%, 4.6%, and 6.6%, respectively. Further information can be found in **Supplementary Table 2**, including the estimated rates of overall and cause-specific mortality, as well as details on individual causes of death. **Figure 4** shows the cause-specific mortality rates over time as a function of CHD severity classification, demonstrating an increased long-term risk of CV mortality for patients with severe CHD that was two- to three-fold higher than those with moderate or mild CHD (e.g., 15-year cardiovascular death rate, 11.0% versus 5.2% or 4.0%, respectively). On the other hand, the comparison of non-CV mortality curves by CHD complexity yielded much smaller differences (15-year non-cardiovascular death rate for severe, moderate, and mild CHD: 6.1%, 3.7%, and 5.6%, respectively).

**Figure 4.**
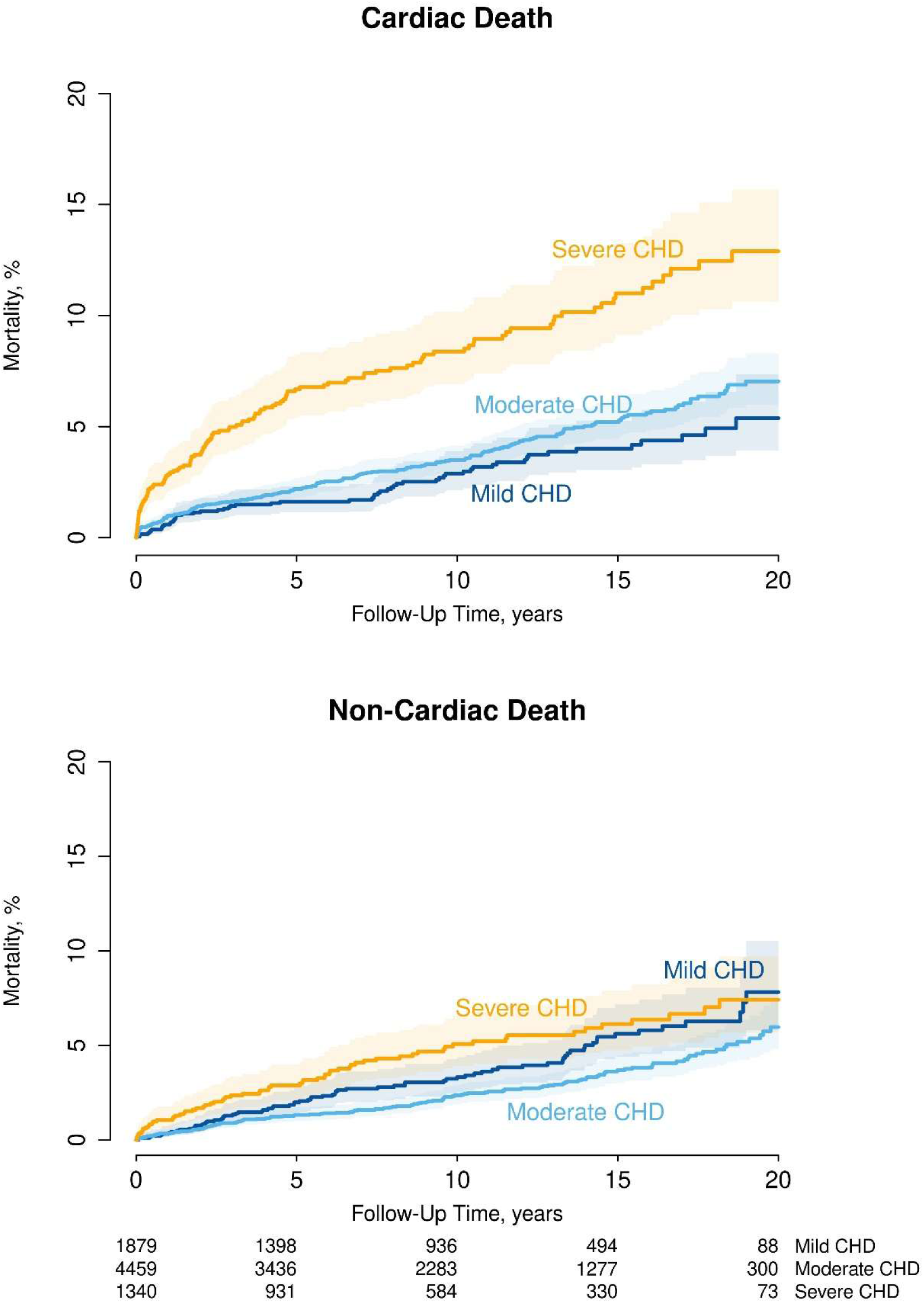
Cumulative incidence curves for cardiac mortality and non-cardiac mortality accounting for the competing risks of non-cardiac and cardiac mortality, respectively, as well as unknown causes of death. The curves are stratified by ACHD severity.

Amongst those who had died from cardiovascular causes, heart failure (48.9%), sudden cardiac death (21.9%) and post-cardiac surgery (13.5%) were the leading causes. The distribution of cardiac-specific causes of death stratified by ACHD severity is shown in **Figure 5A**.

**Figure 5.**
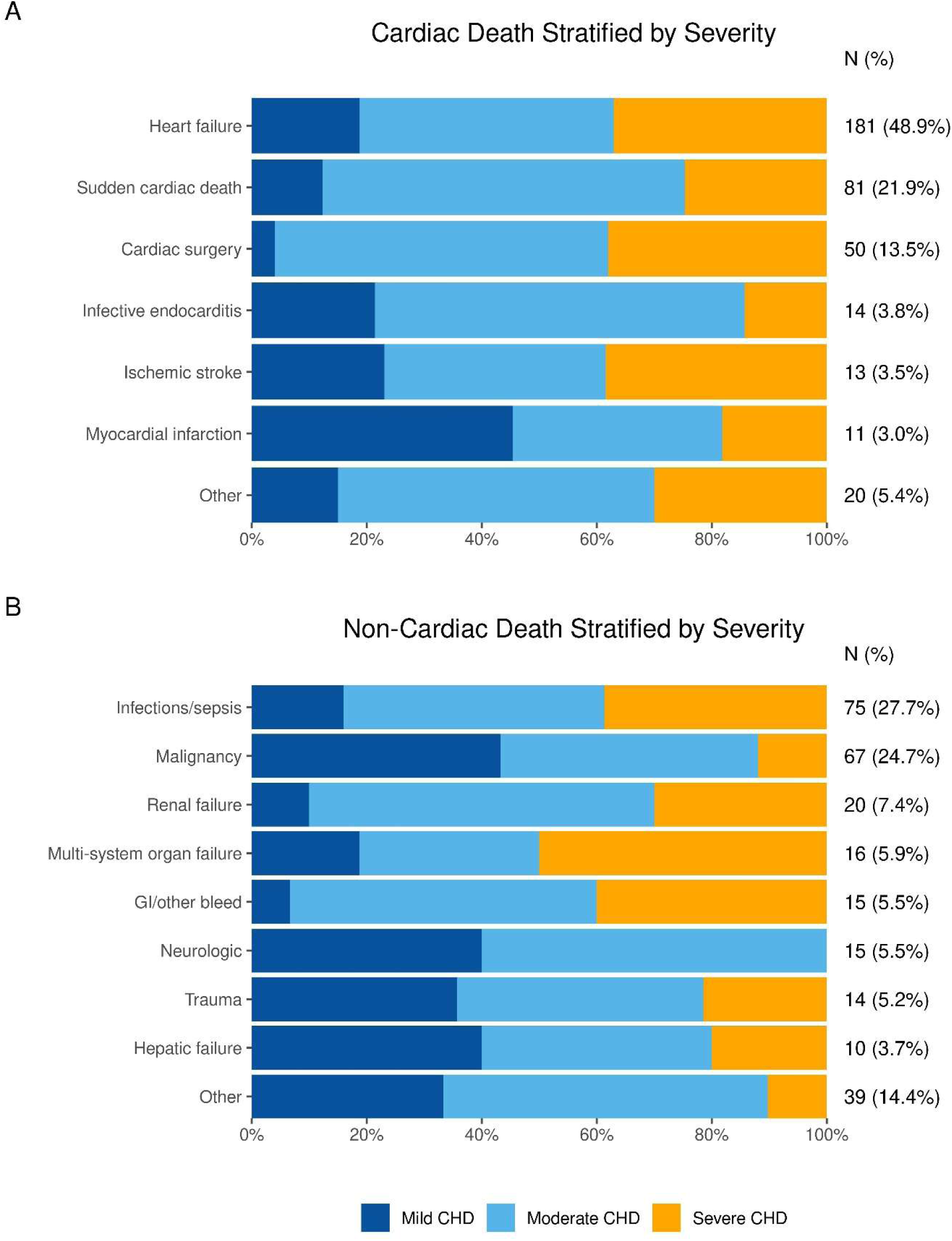
(A) Distribution of cardiac causes of death by ACHD severity. Stacked bar chart showing the proportion of each cardiac cause of death among patients with mild, moderate, and severe ACHD. Heart failure was the leading cardiac cause of death (48.9%), followed by sudden cardiac death (21.9%) and cardiac surgery deaths (13.5%). **(B)** Distribution of non-cardiac causes of death by ACHD severity. *Footnote:* Stacked bar chart showing the proportion of each non-cardiac cause of death among patients with mild, moderate, and severe ACHD. Infection/sepsis (27.7%) and malignancy (24.7%) were the most common non-cardiac causes of death. Note: cardiac surgery includes operative and postoperative deaths. Other cardiac causes of death include heart transplant rejection, vascular rupture/dissection, pulmonary embolism, or hemorrhagic stroke. Other non-cardiac causes of death include lung disease/respiratory failure, neurologic causes, non-cardiac surgery, and suicide/homicide.

Multivariable analysis of risk factors associated with time until cardiovascular death included advanced age, cyanotic heart disease, Fontan palliation, prior valve replacement, significant valve regurgitation, systemic and sub-pulmonary ventricular dysfunction, prior heart failure hospitalization, and NYHA functional class **(Table 2)**. Hypertension, cirrhosis, elevated creatinine levels, and lower hemoglobin levels were additionally associated with increased hazard of cardiovascular death. When the analysis was repeated after excluding the patients who died from unknown causes, the results did not differ appreciably from those in the main analysis censoring those patients **(Supplementary Table 3**).

We further evaluated the unadjusted association between a limited set of baseline factors and the time until sudden cardiac death in separate Cox regression models. Male sex, atrial arrhythmia, and moderate or severe CHD were univariately associated with a 2- to 3-fold increased hazard of sudden cardiac death **(Table 2**).

The cause of death from non-cardiovascular conditions stratified by the severity of ACHD is presented in **Figure 5B**. Deaths due to infection / sepsis (27.7%), malignancy (24.7%), and renal failure (77.4%) were most common in this group.

The cause of death in specific ACHD subgroups are presented in **Figure 6**. Heart failure was predominant cause in most subgroups particularly in systemic right ventricle, single ventricle physiology, and cyanotic heart disease. Sudden cardiac death contributed significantly across all lesion categories, especially in conotruncal lesions, left heart lesions, and systemic right ventricle patients. Non-cardiac causes, including infection, malignancy, and renal failure accounted for a considerable proportion of deaths.

**Figure 6.**
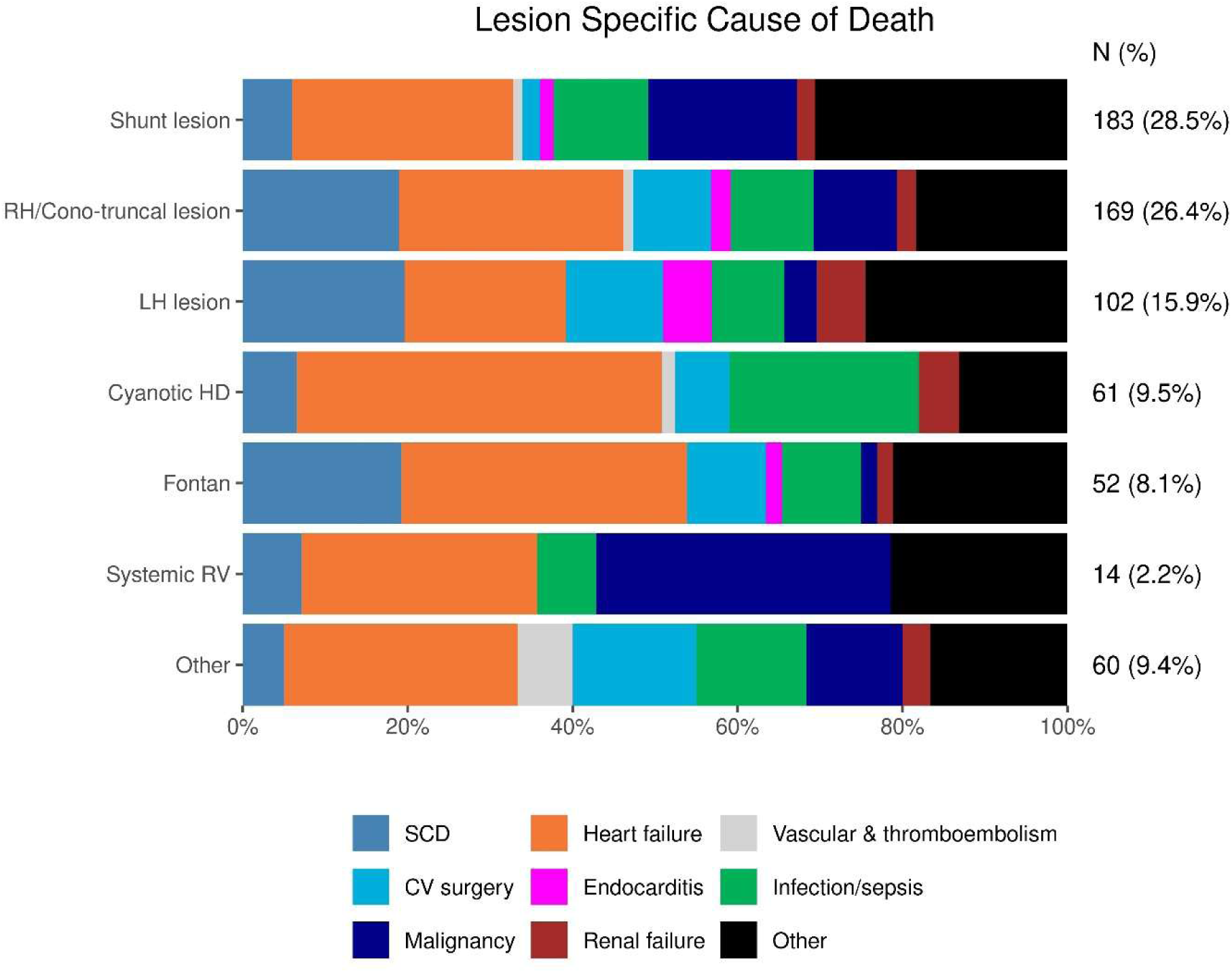
Lesion-specific causes of death in ACHD. Stacked bar chart showing the distribution of major causes of death across congenital heart disease lesion groups. Heart failure, sudden cardiac death, infection, malignancy, and other causes varied by lesion type, highlighting distinct mortality patterns across ACHD subgroups.

## Discussion

In this contemporary cohort of ACHD, we report an annual mortality rate of 1.4%. In addition to age, male sex, and specific subtypes of CHD, risk factors for mortality include ventricular dysfunction and extra-cardiac complications such as renal and liver dysfunction.

Acquired cardiovascular conditions such as hypertension and CAD also emerged as important drivers of mortality reflecting the aging ACHD population. Consistent with prior registry data, heart failure and sudden cardiac death were the leading causes of cardiovascular mortality. In addition to cardiovascular causes, over 40% of known causes of death were non-cardiovascular. These data underscore the need for comprehensive ACHD care models that extend beyond cardiac management.

Our study describes outcomes in a cohort of aging ACHD patients first evaluated at a single center through 2023, adding to previous reports from European centers of patients followed between 2001 and 2015 (4,5). Our annual mortality rate of 1.4 deaths per 100 patient-years, is slightly lower than that reported in the National German Registry (1.7 per 100 patient-years) but higher than the estimate in the Dutch CONCOR registry (0.8 per 100 patient-years) (4,5). Mortality also occurred at a younger age across disease severity in our study compared to previous report from Oliver et al: mild (70.6 vs 78 yrs), moderate (60.2 vs 72.6 yrs), and severe (41.7 vs 49.2 yrs) ACHD groups(6). This may be secondary to the study occurring at a tertiary referral center which may skew towards complex or advanced disease compared to broader population-based registries. These comparisons should therefore be interpreted cautiously given differences in case mix, era, and ascertainment method across registries.

ACHD complexity emerged as an important determinant of risk, with patients with severe lesions—such as Fontan circulation and cyanotic heart disease—demonstrating the highest mortality. However, the mortality rate was not significantly different between mild and moderate ACHD phenotypes, consistent with previous reporting (4,7).

We explore detailed patient level predictors of mortality. In addition to objective echocardiographic measures of systemic and sub-pulmonary ventricular dysfunction, functional measures such as NYHA class and HF related hospitalization were associated with mortality, consistent with previous reports(6,8–10). Consequently, heart failure was the most common cause of mortality. These findings support a management paradigm focused on early recognition and treatment of heart failure.

Sudden cardiac death was the 2^nd^ most common cause of mortality. The risk of sudden cardiac death increased with complexity of ACHD and showed a trend towards association with older age(7,11). The association with atrial arrhythmia and reduced systemic ventricular ejection fraction is consistent with prior reports from smaller CHD cohorts(7,11). These univariable associations need confirmation in larger data sets with multivariable analysis to pave the way for improved SCD risk stratification of ACHD patients in the future.

Hypertension, obesity, smoking, and CAD, were significant associations of mortality. These conditions have previously been reported to be increasingly prevalent in ACHD, highlighting the importance of screening for and early management of these potentially modifiable risk factors in ACHD care(12–14). BMI, another modifiable risk factor, demonstrated a U-shaped relationship with mortality. Underweight patients may represent those with advanced cardiac cachexia, heart failure, or chronic hypoxemia and may identify patients with poor nutritional reserve (15). At the other extreme, BMI≥30kg/m^2^ may increase mortality through the cardiometabolic consequences of excess adiposity as established in the general population (16,17). This bidirectional relationship underscores the importance of nutritional assessment and weight management across the full spectrum of ACHD patients, including those who appear clinically stable.

Abnormal hemoglobin, both anemia and polycythemia, were independently associated with mortality. Prior studies have demonstrated that anemia is prevalent in ACHD and associated with 3-fold increase in mortality (18). The potential role for routine screening for anemia and treatment of reversible causes needs prospective assessment. Likewise polycythemia secondary to cyanotic heart disease is not only associated with advanced disease but also increases risk for thrombotic complications (19).

The improved survival in patients receiving a statin is intriguing and should be considered hypothesis generating. Statin use has been associated with favorable outcomes in various ACHD populations, potentially through pleiotropic anti-inflammatory and endothelial effects in addition to lipid lowering, similar to their established benefit in acquired cardiovascular disease (20,21). Prospective studies to establish indications for and effect of statins in ACHD are warranted. The observed association of loop diuretic, digoxin and reverse association of aspirin with mortality is likely influenced by indication bias and should generally be interpreted with caution.

Non-cardiovascular causes accounted for 42% of deaths with known etiology in our cohort, higher than prior registry estimates (4). This may reflect the evolving demographics of a contemporary ACHD population in whom acquired organ dysfunction, infection and malignancy increasingly compete with cardiac disease as the proximate cause of death. The concentration of infection-related deaths in patients with moderate and severe CHD extended to both infective endocarditis and other infections. Potential etiology include immunological vulnerability, indwelling hardware, residual shunts, and repeated healthcare exposures(22). However, interventions to reduce infections other than infective endocarditis prophylaxis remain poorly defined and in need of further investigation. Malignancy was the second leading non-cardiovascular cause, consistent with reports linking cumulative ionizing radiation exposure from diagnostic and interventional procedures to elevated cancer risk (23). The role of exposure-stratified oncologic surveillance in ACHD warrants further study. Organ failure, predominantly renal and hepatic, may represent end-organ damage from longstanding hemodynamic abnormalities: this cardiorenal and cardiohepatic relationship is well documented in certain ACHD subgroups(24,25). Collectively, these findings support multidisciplinary ACHD care models that formally integrate nephrology, hepatology, infectious disease, and oncology alongside traditional cardiac surveillance.

### Future directions

Several priorities emerge from these findings to guide future investigation. First, prospective studies incorporating structured assessment of physiologic burden including functional capacity, organ reserve, nutritional status, and comorbidity accumulation are needed to determine whether these factors stratify risk beyond anatomic complexity alone and to identify patients who may benefit from earlier or more intensive intervention. Second, the persistently high proportion of sudden cardiac death underscores the urgent need for ACHD specific risk models. Finally, the high burden of non-cardiovascular mortality, particularly from malignancy and infection, calls for prospective evaluation of structured oncologic surveillance protocols and infectious disease co-management pathways in ACHD, neither of which has been formally studied in this population.

### Limitations

This is a retrospective single center study from tertiary care center with potential referral bias. This data is complementary to population registries, and the findings are most applicable to patients seen at ACHD specialty centers. CHD and the cause of death was determined using electronic health record documentation and death certificates; therefore, classification accuracy is dependent on physician documentation and clinical judgment. The cause of death couldn’t be ascertained for 40% of the death cohort. Hence the analysis of causes of death should be considered exploratory.

## Conclusions

In this large contemporary cohort of adults with congenital heart disease followed over two decades, mortality occurred at 1.4% per patient-year. Prognosis was shaped not only by anatomic complexity but by cumulative co-morbidity burden - ventricular dysfunction, heart failure symptoms, acquired cardiovascular risk factors and extra-cardiac organ involvement were all independent predictors of death. Heart failure and sudden cardiac death remained the dominant cardiovascular causes, while non-cardiovascular causes: infection, malignancy, and organ failure accounted for a substantial number of deaths. These findings support a fundamental expansion of the ACHD care model: one that integrates proactive management of acquired cardiovascular risk factors, systematic screening for non-cardiac comorbidities, and multidisciplinary collaboration.

## Data Availability

The data that support the findings of this study are not publicly available because they contain protected patient health information from the Mayo Adult Congenital Heart Disease Registry. De-identified data may be made available from the corresponding author upon reasonable request and with appropriate institutional approvals, subject to Mayo Clinic data-sharing policies.

## Supplementary files

**Supplementary table 1:** Congenital heart diseases included in the cohort.

| **Congenital heart disease diagnosis** |  |
| --- | --- |
| *Right-heart / cono-truncal lesion* |  |
| Tetralogy of Fallot | 1,079 (14.1%) |
| Ebstein anomaly | 744 (9.7%) |
| Pulmonary stenosis or atresia with VSD | 375 (4.9%) |
| Double Outlet Right Ventricle | 46 (0.6%) |
| Truncus Arteriosus | 41 (0.5%) |
| Pulmonary atresia with intact ventricular septum | 50 (0.7%) |
| D-transposition of great arteries with arterial switch | 99 (1.3%) |
| D-transposition of great arteries with Rastelli repair | 31 (0.4%) |
| Double outlet right ventricle with Rastelli repair | 40 (0.5%) |
| *Left heart lesion* |  |
| Coarctation of aorta | 647 (8.4%) |
| Aortic stenosis | 789 (10.3%) |
| Subaortic stenosis | 195 (2.5%) |
| Mitral stenosis | 18 (0.2%) |
| Supravalvular aortic stenosis | 20 (0.3%) |
| *Shunt lesion* |  |
| Atrial septal defect | 904 (11.8%) |
| Partial anomalous pulmonary venous return | 536 (7.0%) |
| Ventricular septal defect (VSD) | 483 (6.3%) |
| Atrioventricular canal defect | 349 (4.5%) |
| *Cyanotic heart disease* |  |
| Unrepaired/palliated Single Ventricle | 86 (1.1%) |
| Eisenmenger syndrome | 100 (1.3%) |
| *Systemic right ventricle* |  |
| Congenitally corrected transposition of great arteries | 246 (3.2%) |
| D-transposition of great arteries with atrial switch | 225 (2.9%) |
| *Single ventricle* |  |
| Fontan palliation | 423 (5.5%) |
| *Other* |  |
| Patent Ductus Arteriosus | 115(1.5%) |
| Cor triatriatum | 37 (0.5%) |

**Supplementary Table 2:** Outcomes table.

| Variable | N | Overall (N=7678) |
| --- | --- | --- |
| All-Cause Mortality | 7678 |  |
| Events |  | 1116 |
| years = 5 |  | 6.9% (493) |
| years = 10 |  | 12.0% (755) |
| years = 15 |  | 19.0% (996) |
| years = 20 |  | 26.2% (1109) |
| # events/patient-years (%) |  | 1116/78767.8 (1.4%) |
| Cause of death | 1116 |  |
| CV-related |  | 370 (33.2%) |
| Non-CV related |  | 271 (24.3%) |
| Unknown |  | 475 (42.6%) |
| CV-related death<sup>1</sup> | 7678 |  |
| Events |  | 370 |
| years = 5 |  | 2.8% (204) |
| years = 10 |  | 4.2% (275) |
| years = 15 |  | 5.9% (336) |
| years = 20 |  | 7.6% (366) |
| Sudden cardiac death<sup>2</sup> | 7678 |  |
| Events |  | 81 |
| years = 5 |  | 0.6% (41) |
| years = 10 |  | 0.9% (59) |
| years = 15 |  | 1.3% (71) |
| years = 20 |  | 1.9% (81) |
| Specific cause of CV-related death: | 370 |  |
| Sudden cardiac death |  | 81 (21.9%) |
| Heart failure |  | 181 (48.9%) |
| Stroke/systemic embolism |  | 13 (3.5%) |
| Hemorrhagic stroke/intracranial hemorrhage |  | 8 (2.2%) |
| Pulmonary Embolism |  | 4 (1.1%) |
| Vascular rupture/dissection |  | 5 (1.4%) |
| Endocarditis |  | 14 (3.8%) |
| Cardiac surgery |  | 50 (13.5%) |
| Myocardial infarction |  | 11 (3.0%) |
| Heart transplant rejection |  | 3 (0.8%) |
| Non-CV related death<sup>3</sup> | 7678 |  |
| Events |  | 271 |
| years = 5 |  | 1.8% (125) |
| years = 10 |  | 3.1% (192) |
| years = 15 |  | 4.6% (242) |
| years = 20 |  | 6.6% (271) |
| Specific cause of non-CV related death: | 271 |  |
| Multisystem organ failure* |  | 16 (5.9%) |
| Malignancy |  | 67 (24.7%) |
| Renal failure |  | 20 (7.4%) |
| Infection/Sepsis (other than endocarditis) |  | 75 (27.7%) |
| GI bleeding/other major bleeding |  | 15 (5.5%) |
| Neurologic cause |  | 15 (5.5%) |
| Trauma/motor vehicle accident |  | 14 (5.2%) |
| Hepatic failure |  | 10 (3.7%) |
| Post surgery (within 30 days) |  | 6 (2.2%) |
| Suicide/homicide |  | 2 (0.7%) |
| COPD exacerbation |  | 7 (2.6%) |
| Restrictive lung disease |  | 5 (1.8%) |
| Respiratory failure |  | 4 (1.5%) |
| Other |  | 15 (5.5%) |
| *Multisystem organ failure is defined as dysfunction of $\geq 2$ organ systems requiring medical intervention to maintain the homeostasis. | | |

Values represent cumulative incidence percent (cumulative number of events) for time-to-event variables and frequency (percentage) for categorical variables. N is the number of non-missing values.
<sup>1</sup> Accounting for the competing risks of non-CV related death and unknown CoD.
<sup>2</sup> Accounting for the competing risks of other CV-related death (non-SCD), non-CV related death, and unknown CoD.
<sup>3</sup> Accounting for the competing risk of CV related death and unknown CoD.

**Supplementary Table 3:** Sensitivity multivariable analysis for cardiovascular death excluding patients who died due to unknown causes.

| Variable | cHR (95% CI) | P |
| --- | --- | --- |
| Age, years |  | <0.001 |
| 35 (reference) | 1.0 |  |
| 50 | 1.8 (1.6 - 2.0) |  |
| 65 | 4.2 (3.2 - 5.5) |  |
| Male gender | 1.1 (0.8 - 1.4) | 0.528 |
| CHD group |  | <0.001 |
| Right heart /Cono-truncal lesion (reference) | 1.0 |  |
| Left heart lesion | 1.4 (1.0 - 2.0) |  |
| Shunt lesion | 0.9 (0.6 - 1.3) |  |
| Cyanotic heart disease | 5.0 (3.2 - 8.0) |  |
| Systemic right ventricle | 0.8 (0.5 - 1.3) |  |
| Fontan palliation | 2.7 (1.6 - 4.6) |  |
| Other CHD with biventricular repair and systemic LV | 1.1 (0.5 - 2.6) |  |
| Valve replacement | 1.4 (1.1 - 1.9) | 0.011 |
| Prior palliative shunt | 1.2 (0.9 - 1.6) | 0.317 |
| Hypertension | 1.3 (1.0 - 1.7) | 0.043 |
| Hyperlipidemia | 0.6 (0.4 - 0.8) | 0.002 |
| Smoking history | 1.1 (0.9 - 1.5) | 0.318 |
| Coronary artery disease | 1.4 (1.0 - 2.1) | 0.049 |
| Atrial arrhythmia | 1.3 (1.0 - 1.6) | 0.058 |
| Ventricular arrhythmia | 1.4 (1.0 - 2.0) | 0.091 |
| Infective endocarditis | 1.4 (1.0 - 2.0) | 0.080 |
| Cirrhosis | 1.8 (1.1 - 3.1) | 0.031 |
| HF hospitalization | 2.2 (1.7 - 2.9) | <0.001 |
| New York Heart Association functional class | 1.2 (1.1 - 1.4) | 0.004 |
| Body mass index, kg/m<sup>2</sup> |  | 0.288 |
| 20 | 0.9 (0.7 - 1.1) |  |
| 25 (reference) | 1.0 |  |
| 30 | 1.1 (1.0 - 1.2) |  |
| 40 | 1.2 (0.8 - 1.8) |  |
| Creatinine, mg/dL |  | <0.001 |
| 0.6 | 0.9 (0.6 - 1.3) |  |
| 0.8 (reference) | 1.0 |  |
| 1 | 1.2 (1.1 - 1.3) |  |
| 1.3 | 1.5 (1.4 - 1.7) |  |
| Hemoglobin, g/dL |  | 0.003 |
| 17 | 1.1 (0.9 - 1.4) |  |
| 15 (reference) | 1.0 |  |
| 13 | 1.0 (0.9 - 1.1) |  |
| 11 | 1.3 (1.1 - 1.6) |  |
| 9 | 1.8 (1.3 - 2.6) |  |
| *Echocardiographic parameters* |  |  |
| Systemic ventricular ejection fraction |  | 0.007 |
| 60 (reference) | 1.0 |  |
| 45 | 1.4 (1.1 - 1.8) |  |
| 30 | 1.9 (1.2 - 3.1) |  |
| ≥Moderate RV dysfunction | 1.5 (1.0 - 2.3) | 0.057 |
| ≥Moderate valve regurgitation | 1.4 (1.1 - 1.8) | 0.008 |
| *Medications* |  |  |
| Loop diuretics | 1.4 (1.1 - 1.8) | 0.011 |
| Beta blockers | 0.8 (0.7 - 1.1) | 0.121 |
| Angiotensin converting enzyme inhibitor / aldosterone receptor blocker | 1.2 (0.9 - 1.5) | 0.257 |
| Aspirin | 0.6 (0.5 - 0.8) | <0.001 |
| Anti-arrhythmic drug | 0.8 (0.6 - 1.1) | 0.165 |
| Digoxin | 1.5 (1.1 - 2.0) | 0.007 |
| Statin | 0.6 (0.4 - 0.8) | 0.003 |

cHR: cause-specific hazard ratio (censoring lost to follow-up and death due to non-cardiovascular causes); CI: confidence interval.

**Supplementary Table 4:**
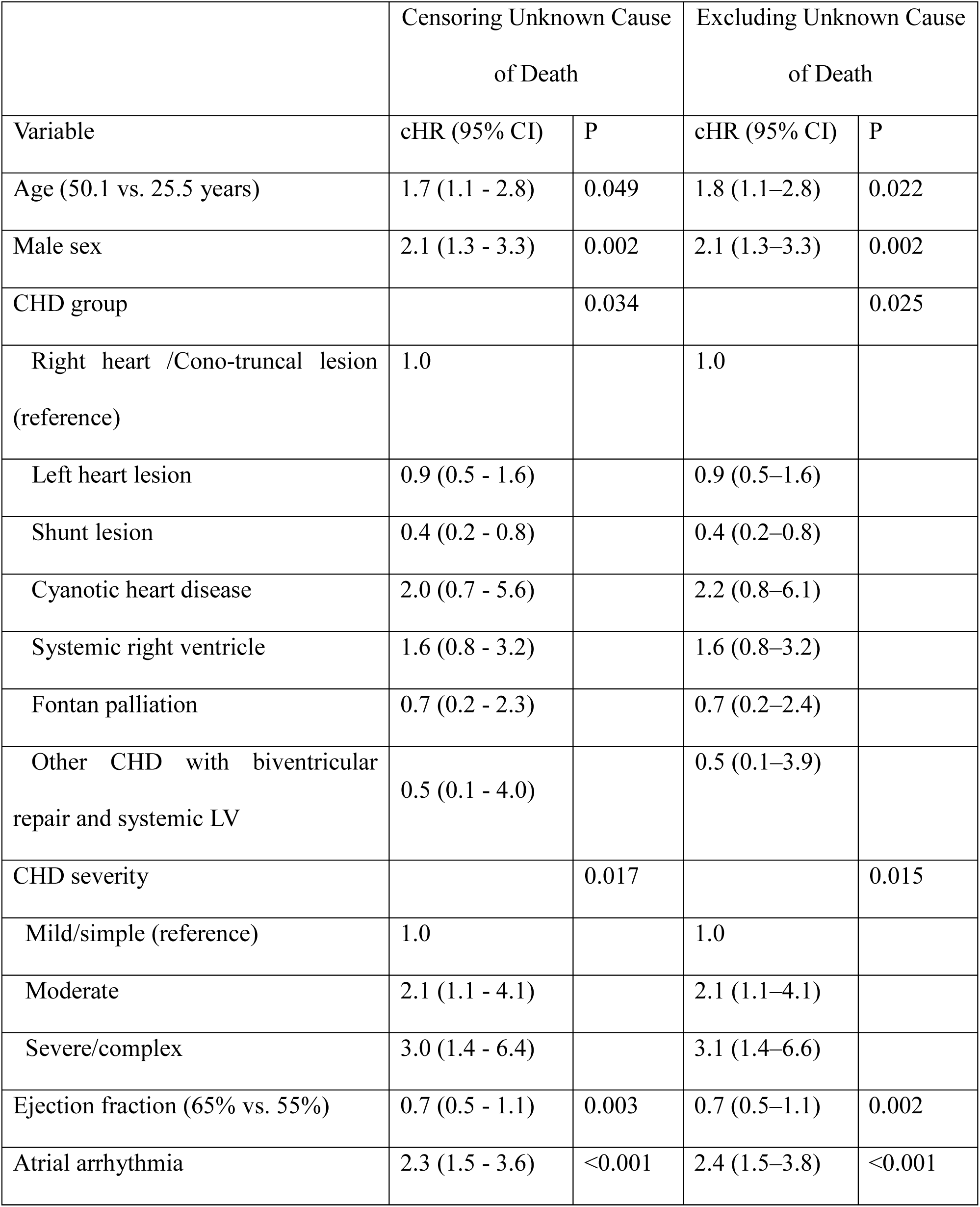
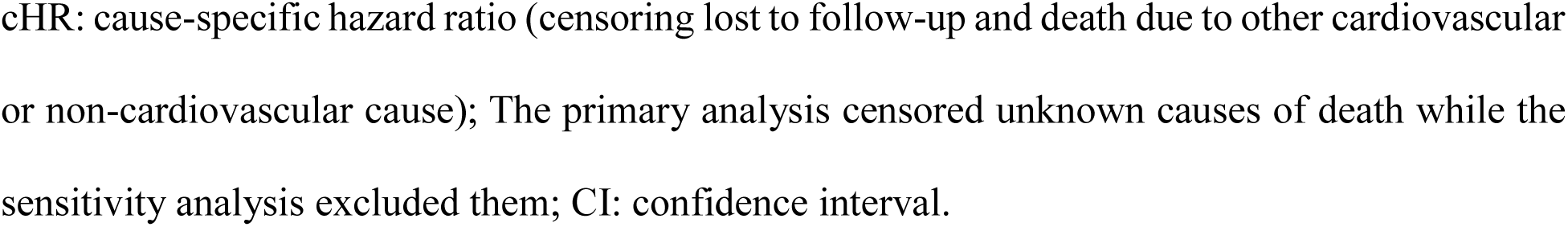
Univariable analysis for sudden cardiac death.

| Variable | Censoring Unknown Cause<br>of Death |  | Excluding Unknown Cause<br>of Death |  |
| --- | --- | --- | --- | --- |
|  | cHR (95% CI) | P | cHR (95% CI) | P |
| Age (50.1 vs. 25.5 years) | 1.7 (1.1 - 2.8) | 0.049 | 1.8 (1.1–2.8) | 0.022 |
| Male sex | 2.1 (1.3 - 3.3) | 0.002 | 2.1 (1.3–3.3) | 0.002 |
| CHD group |  | 0.034 |  | 0.025 |
| Right heart /Cono-truncal lesion<br>(reference) | 1.0 |  | 1.0 |  |
| Left heart lesion | 0.9 (0.5 - 1.6) |  | 0.9 (0.5–1.6) |  |
| Shunt lesion | 0.4 (0.2 - 0.8) |  | 0.4 (0.2–0.8) |  |
| Cyanotic heart disease | 2.0 (0.7 - 5.6) |  | 2.2 (0.8–6.1) |  |
| Systemic right ventricle | 1.6 (0.8 - 3.2) |  | 1.6 (0.8–3.2) |  |
| Fontan palliation | 0.7 (0.2 - 2.3) |  | 0.7 (0.2–2.4) |  |
| Other CHD with biventricular<br>repair and systemic LV | 0.5 (0.1 - 4.0) |  | 0.5 (0.1–3.9) |  |
| CHD severity |  | 0.017 |  | 0.015 |
| Mild/simple (reference) | 1.0 |  | 1.0 |  |
| Moderate | 2.1 (1.1 - 4.1) |  | 2.1 (1.1–4.1) |  |
| Severe/complex | 3.0 (1.4 - 6.4) |  | 3.1 (1.4–6.6) |  |
| Ejection fraction (65% vs. 55%) | 0.7 (0.5 - 1.1) | 0.003 | 0.7 (0.5–1.1) | 0.002 |
| Atrial arrhythmia | 2.3 (1.5 - 3.6) | <0.001 | 2.4 (1.5–3.8) | <0.001 |

cHR: cause-specific hazard ratio (censoring lost to follow-up and death due to other cardiovascular or non-cardiovascular cause); The primary analysis censored unknown causes of death while the sensitivity analysis excluded them; CI: confidence interval.

## References

1. Shekhar S, Agrawal A, Pampori A, Lak H, Windsor J, Ramakrishna H. Mortality in Adult Congenital Heart Disease: Analysis of Outcomes and Risk Stratification. J Cardiothorac Vasc Anesth 2022;36:3379–3388.

2. Müller MJ, Norozi K, Caroline J et al. Morbidity and mortality in adults with congenital heart defects in the third and fourth life decade. Clin Res Cardiol 2022;111:900–911.

3. Warnes CA, Liberthson R, Danielson GK et al. Task force 1: the changing profile of congenital heart disease in adult life. J Am Coll Cardiol 2001;37:1170–5.

4. Verheugt CL, Uiterwaal CS, van der Velde ET et al. Mortality in adult congenital heart disease. Eur Heart J 2010;31:1220–9.

5. Engelings CC, Helm PC, Abdul-Khaliq H et al. Cause of death in adults with congenital heart disease - An analysis of the German National Register for Congenital Heart Defects. Int J Cardiol 2016;211:31–6.

6. Oliver JM, Gallego P, Gonzalez AE et al. Risk factors for excess mortality in adults with congenital heart diseases. Eur Heart J 2017;38:1233–1241.

7. Vehmeijer JT, Koyak Z, Leerink JM et al. Identification of patients at risk of sudden cardiac death in congenital heart disease: The PRospEctiVE study on implaNTable cardIOverter defibrillator therapy and suddeN cardiac death in Adults with Congenital Heart Disease (PREVENTION-ACHD). Heart Rhythm 2021;18:785–792.

8. Ombelet F, Goossens E, Apers S, Budts W, Gewillig M, Moons P. Predicting 15-Year Mortality in Adults With Congenital Heart Disease Using Disease Severity and Functional Indices. Can J Cardiol 2019;35:907–913.

9. Trojnarska O, Grajek S, Katarzyński S, Kramer L. Predictors of mortality in adult patients with congenital heart disease. Cardiol J 2009;16:341–7.

10. Zomer AC, Vaartjes I, van der Velde ET et al. Heart failure admissions in adults with congenital heart disease; risk factors and prognosis. International Journal of Cardiology 2013;168:2487–2493.

11. Koyak Z, Harris L, de Groot JR et al. Sudden cardiac death in adult congenital heart disease. Circulation 2012;126:1944–54.

12. Bhatt AB, Foster E, Kuehl K et al. Congenital heart disease in the older adult: a scientific statement from the American Heart Association. Circulation 2015;131:1884–931.

13. Brida M, De Rosa S, Legendre A et al. Acquired cardiovascular disease in adults with congenital heart disease. Eur Heart J 2023;44:4533–4548.

14. Engelfriet PM, Drenthen W, Pieper PG et al. Smoking and its effects on mortality in adults with congenital heart disease. Int J Cardiol 2008;127:93–7.

15. Vest AR, Chan M, Deswal A, et al. Nutrition, Obesity, and Cachexia in Patients With Heart Failure: A Consensus Statement from the Heart Failure Society of America Scientific Statements Committee. J Card Fail 2019;25:380–400.

16. Iyen B, Weng S, Vinogradova Y, Akyea RK, Qureshi N, Kai J. Long-term body mass index changes in overweight and obese adults and the risk of heart failure, cardiovascular disease and mortality: a cohort study of over 260,000 adults in the UK. BMC Public Health 2021;21:576.

17. Andonian C, Langer F, Beckmann J et al. Overweight and obesity: an emerging problem in patients with congenital heart disease. Cardiovasc Diagn Ther 2019;9:S360–s368.

18. Dimopoulos K, Diller GP, Giannakoulas G et al. Anemia in adults with congenital heart disease relates to adverse outcome. J Am Coll Cardiol 2009;54:2093–100.

19. Perloff JK, Rosove MH, Child JS, Wright GB. Adults with cyanotic congenital heart disease: hematologic management. Ann Intern Med 1988;109:406–13.

20. Flannery LD, Fahed AC, DeFaria Yeh D et al. Frequency of Guideline-Based Statin Therapy in Adults With Congenital Heart Disease. Am J Cardiol 2018;121:485–490.

21. Lee SE, Chang HJ, Sung JM et al. Effects of Statins on Coronary Atherosclerotic Plaques: The PARADIGM Study. JACC Cardiovasc Imaging 2018;11:1475–1484.

22. Kuijpers JM, Koolbergen DR, Groenink M et al. Incidence, risk factors, and predictors of infective endocarditis in adult congenital heart disease: focus on the use of prosthetic material. Eur Heart J 2017;38:2048–2056.

23. Cohen S, Liu A, Gurvitz M et al. Exposure to Low-Dose Ionizing Radiation From Cardiac Procedures and Malignancy Risk in Adults With Congenital Heart Disease. Circulation 2018;137:1334–1345.

24. Jang SY, Kim EK, Chang SA et al. Prognosis of Chronic Kidney Disease and Metabolic Syndrome in Adults With Congenital Heart Disease. J Korean Med Sci 2023;38:e375.

25. Cao JY, Wales KM, d’Udekem Y, Celermajer DS, Cordina R, Majumdar A. Prevalence, Risk Factors, and Prognosis for Fontan-Associated Liver Disease: A Systematic Review and Exploratory Meta-Analysis. JACC Adv 2025;4:101694.

